# Evaluation of trace elements analysis in Pediatric Patients with COVID-19: A report from Turkey

**DOI:** 10.1101/2022.07.20.22277852

**Authors:** Tahir Dalkıran, Velid Unsal, Sevcan İpek, Dogan Oncu, Mehmet Mercan, Yaşar Kandur

**Author notes:** **Corresponding author:** Yasar Kandur,M.D., Department of Pediatrics, Faculty of Medicine, Kırıkkale University, Kırıkkale, Turkey, <u>Tel:90</u> 3183574242.

## Abstract

**Introduction:** This study aimed to evaluate the levels of Selenium, Copper, and Zinc in an attempt to identify the role of trace elements in pediatric patients with COVID-19 infection.

**Methods:** We randomly selected 29 patients who were hospitalized with the diagnosis of COVID-19. Blood serum sample was collected to study serum Se, Zn, and Cu levels at disease onset and at the time of discharge.

**Results:** The median age of our patients was 71.1 ± 14.4 months (range, 3-205 months); 14 (48.3%) patients were male. The mean CRP, and D-Dimer levels were significantly higher at disease onset than at discharge. On the other hand, the mean Cu, Zn, and Se levels were significantly lower at disease onset compared to the time of discharge. The patients with mild-moderate disease severity were older than the patients with severe disease although the difference did not reach statistical significance (82.2±17.7 vs 54.5± 24.7 months; p=0.374). There was no correlation between age and trace elements other than Zn.

**Conclusion:** We believe that, patients and other individuals under risk of COVID-19 should be supplemented with trace elements.

## Introduction

Coronavirus disease 2019 (COVID-19) has become a public health threat to people all over the world during the last 10 months. The lower airway is the primary target of the infection.^1^ Acute respiratory distress syndrome (ARDS), septic shock, and coagulation disorders are severe complications of this infection, but such severity is rare in children.^2^ Trace elements are a significant factor for the immune system, as such studies have now established a significant association between trace elements, such as selenium (Se), Copper (Cu), Zinc (Zn), and COVID-19 prognosis. Specific selenoproteins especially selenocysteine that have a role in immune system need abundant Se supply for their enzymatic activity.^3^ As such, Se deficiency is an established risk factor for viral infections.^4^ Another trace element, Zn, is an essential component of superoxide dismutase 1 and 3,^5^ and is an important factor for the development and maintenance of immune system.^6^ Hence, Zn deficiency is known to result in humoral and cell-mediated immunity dysfunction.^7^ Cu has been associated with IL-2 production in immune response.^8^ Cu is further involved in T cell proliferation,^9^ antibody production, and cellular immunity.^10^ This study aimed to evaluate the levels of Se, Cu, and Zn in an attempt to identify the role of trace elements in pediatric patients with COVID-19 infection.

## Methods

We randomly selected 29 patients who were hospitalized with the diagnosis of COVID-19 during a period between May 25 to August 12, 2020, at a local state and university hospital. Written informed consent was obtained from the patients’ parents. Data on demographic properties, medical history, symptoms, signs, and laboratory findings were collected from the patients’ medical records. Blood serum sample was collected to study serum Se, Zn, and Cu levels at disease onset and at the time of discharge.

For the trace element analysis, Bruker Icp MS Aurora M90 model was used. Five ml of nitric acid and 5 ml of ultrapure water were added on it. The solution mixture was mixed. Ten minutes was waited for preburning. Later, the tubes were covered and fired in a microwave oven. The burning program in a microwave oven (human blood): Initially, the solution was microwaved for 20 min, and it was heated up to 200°C throughout. Then, it was kept at 200°C for 15 minutes. Pressure was set at 800 psi, and power at 900-1800 watts. Then,it was cooled from 200°C to room temperature within 15 minutes. The samples taken from the tubes after burning were completed up to 50 ml with ultrapure water. The results were expressed in µg/L.

### Statistical Analysis

The categorical variables were expressed as frequency and percentage, and continuous variables as mean and standard deviation (SD). Independent samples t test was used for the comparison of means of normally distributed continuous variables; Mann-Whitney U test was used for non-normally distributed continuous variables. All statistical analyses were performed using SPSS (Statistical Package for the Social Sciences) version 20.0 software (SPSS Inc.). Two-sided P-values of less than 0.05 were considered statistically significant. Correlation analysis between the laboratory variables and the cytokines was performed using a non-parametric test (Spearman rank correlation test). Ethics committee approval was obtained from the Kahramanmaras Sütçü Imam University Clinical Research Ethics Committee (Date: 27.5.2020, No:2020/10).

## Results

The median age of our patients was 71.1 ± 14.4 months (range, 3-205 months); 14 (48.3%) patients were male. Eleven (37.9%) patients had a history of contact with a COVID-positive person. The presenting symptoms were were as follows; fever (n=20, 69%), cough (n=15, 51.7%), respiratory distress (n=12, 41.4%), nasal congestion/runny nose (n=10, 34.4%), headache (n=2, 6.8%), and sore throat (n=2, 6.8 %). One patient had also abdominal pain. None of the patients had myalgia. Physical examination signs were as follows; 13 (44.8%) patients had rales/ronchi on lung auscultation, 4 (13.48%) patients had cyanosis at hospital admission,11 (37.9%) patients had wheezing, 9 (31.0%) patients had signs and symptoms of respiratory distress (dyspnea, suprasternal and intercostal retractions). Sixteen (55.1%) patients had tachycardia and tachypnea. Two (6.8%) patients were asymptomatic. Ten (34.4%) patients had severe disease and were admitted to the intensive care unit. The remaining 19 patients had mild-moderate disease. None of the patients died from the disease.

The mean CRP, and D-Dimer levels were significantly higher at disease onset than at discharge. On the other hand, the mean Cu, Zn, and Se levels were significantly lower at disease onset compared to the time of discharge (Table 1).There was no difference between the two genders with espect to mean Cu, Zn; however, the mean Se level was significantly higher in males compared to females (44±13 vs 30±15 µg/L; p=0.034) (Table 2).

**Table 1.**
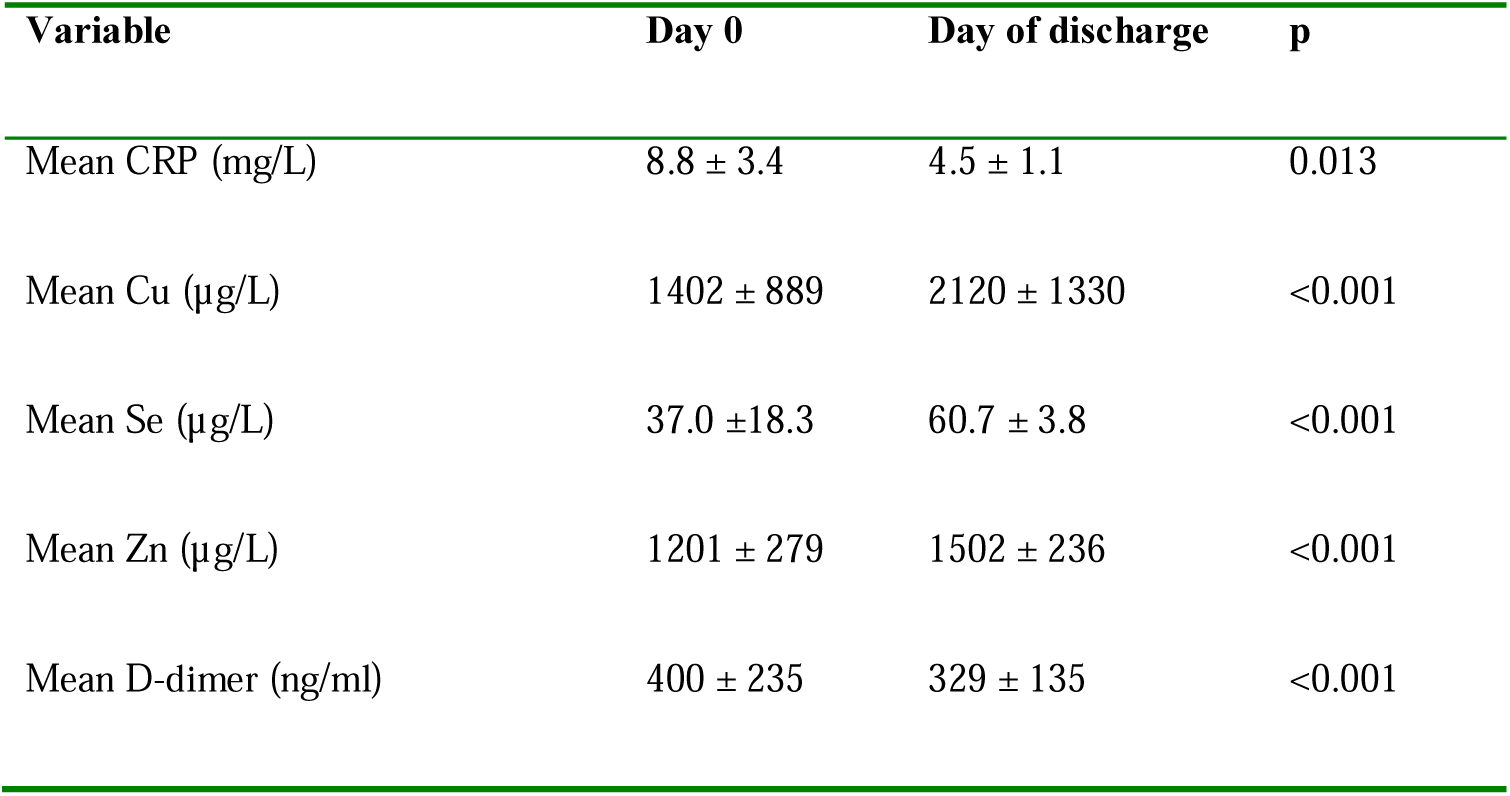
Comparison of the antioxidative markers and trace elements taken at different times of the disease course.

**Table 2.** Comparison of genders with respect to mean antioxidative marker levels and trace elements.

| **Variable** | **Male** | **Female** | **p** |
| --- | --- | --- | --- |
| Mean Cu (µg/L) | 1617±1170 | 1202±465 | 0.215 |
| **Mean Se (µg/L)** | **44±13** | **30±15** | **0.034** |
| Mean Zn (µg/L) | 1219±228 | 1196±328 | 0.833 |
| Mean D-dimer (ng/ml) | 446±266 | 362±212 | 0.445 |

The patients with mild-moderate disease severity were older than the patients with severe disease although the difference did not reach statistical significance (82.2±17.7 vs 54.5± 24.7 months; p=0.374 There was no significant difference between the disease clinics with regard to the levels of trace elements other than Se. The mean Se level at the time of discharge were significantly higher in patients with severe disease.

Six (17.1%) patients had wide spread groundglass opacity (GGO) and multifocal consolidations on thoracic computed tomography. Three patients needed nasal CPAP therapy. Two patients needed mechanical ventilation.

Correlation analyses showed a positive correlation between age and Zn levels (disease onset r=0.387; p=0.05, and recovery r=0.487; p= 0.01). There was no correlation between age and trace elements other than Zn. There was no corellation between CRP and trace elements,

Regression analyses showed that, age, D-dimer, Zn, Se, Cu, are not risk factors for disease severity whereas high CRP (OR=0.282, 95% CI: 0.099-0.557; p=0.007) was a risk factors for severe disease and poor prognosis.

## Discussion

Although there are letters and reviews that reveal the relationship between the trace elements and COVID-19 severity so far, as far as we know there are no published peer-reviewed research in pediatric age group. The evidence that the the trace elements Zn, Se, and Cu involved in the course and outcome of the COVID-19 disease is observational and weak. However, in cases of severe acute respiratory syndrome coronavirus (SARS) and other viral infections, it was observed that supplements administered at an early stage support host resistance against RNA viral infections, which might also include severe COVID-19.^11^

Copper, Zn, and Se provide an antioxidant function with very different mechanisms. Selenium is an essential trace element with antioxidant, immunological, and anti-inflammatory properties.^12,13^ Copper and Zn participate in the enzymatic mechanisms of superoxide dismutase (SOD), which protect against free radicals, and therefore, play an important auxiliary role in oxidative balance.^14^

Zinc is one of the main factors controlling the function and proliferation of neutrophils, natural killer cells, macrophages and T and B lymphocytes, as well as cytokine production by immune cells. Zn also mediates protection from the negative impact of ROS, which is usually produced during inflammatory processes.^15^ Zinc deficiency has been reported in critically ill patients with septic shock.^16^ Lee et al. included a large number of patients with septic disease and showed that 86-95% of critically ill patients had low Zn levels.^17^ It has been suggested that SARS-CoV-2 interacts with angiotensin-converting enzyme 2 (ACE2) in the alveoli which is a zinc-dependent enzyme.^18^ Recent studies showed that a reduced Zn level favors the interaction between ACE2 and SARS-CoV-2 spike protein. Hence, an increased Zn level inhibits ACE2 expression, resulting in reduced viral interaction.^19,20.^ In this manuscript, we report that patients had a low level of zinc at disease onset compared to its recovery phase. However, there was no difference between severe and non-severe patients in mean of zinc level. We also found that low Zn was a not a risk a factor for severe disease. However, probably a low Zn level may a pose risk for contracting the disease. Therefore, it may be beneficial to take Zn during this pandemic.

Selenium is an integral part of Se dependent Glutathione peroxidase (GSH-px), which catalyzes the degredation of H2O2, a ROS. The activity of Se-GSH-px decreases in dietary Se deficiency.^21^ There was no significant difference in terms of mean Se levels between severe and non-severe disease, except for a higher level of Se level at discharge, which was lower in severe patients at discharge.We thougt that the Se is consumed more in severe disease. In disease and in the case of worsening inflammation, a potentially pre-existing low Se level may decline further. This finding is supported by similar findings in other severe diseases, such as sepsis^22^ and polytraumatic injury,^23^ where low, declining, and mortality-relevant Se deficiency has been observed that is unlikely a predisposition. Secondly, a longer stay at the ICU under inflammatory and hypoxic conditions may cause an elevated Se requirement due to ongoing Se loss.^24^ Our population composed of a high proportion of severe patients compared to the normal population, leading to a finding of low Se level in the recovery period. In this pandemic, it seems reasonable to recommend Se, especially for patients at risk.

Copper has been shown to have a role in the innate immune response.^25^ It has been associated with IL-2 production, T cell proliferation,^26^ antibody production, and cellular immunity.^27^ The copper levels were lower at disease onset than recovery, but there was no difference between severe and non-severe cases. Copper has the capability of destroying viruses by contact killing, including SARS-CoV-2. In a cell-based study, Cu was shown to block papain-like protease-2, a protein that SARS-CoV-1 needs for replication.^27^ Low Cu levels suggest a role in the course of the disease and superoxide activity. We thougt that the copper is consumed more in severe disease as selenium.It is possible that Cu, like in other viral disease, may have blocked the protein that the COVID-19 agent needs for replication. However, this hypothesis can only be elucidated by further research.

Our study has some limitations. First, our patient goup is not homogenous since there are more severe patients than present in previos studies. Secondly, this study only included a small number of patients; thus, the results should be interpreted with caution, and statistical non-significance may not rule out differences between severe and mild cases. Thirdly we did not compared the values with a control group of healthy individuals.

## Data Availability

The data supporting this research article are available from the corresponding author on reasonable request.

## Conclusion

COVID-19 constitutes a universal threat nowadays, necessitating strengthening the immune system. Patients and other individuals under risk of COVID-19 should be supplemented with trace elements. In conclusion, our findings support the idea that trace elements are implicated in the success or failure of the response to virus infection.

## DISCLOSURE

Authors disclose that they have no competing interests.

### Authors’ contributions

TD, YK contributed to analysis and drafted the manuscript. SI,DO and MM helped evaluate subjects. YK design and critically revised the manuscript; VU contributed to conception and design and contributed to interpretation. All authors have read and approved the manuscript.

### Ethical approval

Ethics committee approval was obtained from the Kahramanmaras Sütçü Imam University Clinical Research Ethics Committee (Date: 27.5.2020, No:2020/10).

## References

1. Lai CC, Shih TP, Ko WC, Tang HJ, Hsueh PR. Severe acute respiratory syndrome coronavirus 2 (SARS-CoV-2) and corona virus disease-2019 (COVID-19): the epidemic and the challenges. Int J Antimicrob Agent. 2020; 55:105924.

2. Chen F, Liu ZS, Zhang FR, Xiong RH, Chen Y, Cheng XF, et al. First case of severe childhood novel coronavirus pneumonia in China. Zhonghua Er Ke Za Zhi. 2020; 58:179–182

3. Ho mann PR, Berry MJ. The influence of selenium on immune responses. Mol. Nutr. Food Res. 2008, 52, 1273–1280

4. Guillin OM, Vindry C, Ohlmann T, Chavatte L. Selenium, selenoproteins and viral infection. Nutrients 2019, 11, 2101

5. Tainer JA, Getzo ED, Richardson JS. Richardson, D.C. Structure and mechanism of copper, zinc superoxide dismutase. Nature 1983, 306, 284–287.

6. Maares M, Haase H. Zinc and immunity: An essential interrelation. Arch. Biochem. Biophys 2016, 611, 58–65.

7. Tuerk MJ, Fazel N. Zinc deficiency. Curr. Opin. Gastroenterol. 2009, 25, 136–143.

8. Iddir M, Brito A, Dingeo G, et al. Strengthening the immune system and reducing inflammation and oxidative stress through diet and nutrition: considerations during the COVID-19 crisis. Nutrients 2020;12(6)E1562,

9. Saeed F, Nadeem, M Ahmed RS, Tahir Nadeem M, Arshad M.S, Ullah A. Studying the impact of nutritional immunology underlying the modulation of immune responses by nutritional compounds—A review. Food Agric. Immunol. 2016, 27, 205–229.

10. Wintergerst ES, Maggini S, Hornig DH. Contribution of selected vitamins and trace elements to immune function. Ann. Nutr. Metab. 2007, 51, 301–323.

11. Calder PC, Carr AC, Gombart AF, Eggersdorfer M. Optimal Nutritional Status for aWell-Functioning Immune System Is an Important Factor to Protect against Viral Infections. Nutrients 2020, 12, 1181.

12. Fedor M, Urban B, Socha K, Soroczynska J, Kretowska M, Borawska M H, Bakunowicz-Łazarczyk A. Concentration of Zinc, Copper, Selenium, Manganese, and Cu/Zn Ratio in Hair of Children and Adolescents with Myopia. Journal of ophthalmology. 2019; 2019: 5643848.

13. Rech M, To L, Tovbin A, Smoot T, Mlynarek M. Heavy metal in the intensive care unit: a review of current literature on trace element supplementation in critically ill patients. Nutrition in clinical practice. 2014;29(1), 78–89.

14. Ozcelik D, Ozaras R, Gurel Z, Uzun H, Aydin S. Copper-mediated oxidative stress in rat liver. Biological trace element research. 2003; 96(1-3), 209-215.

15. Rahman MT, Idid SZ. Can Zn Be a Critical Element in COVID-19 Treatment?. Biological Trace Element Research. 2020;1-9.

16. Besecker BY, Exline MC, Hollyfield J, Phillips G, Disilvestro RA, Wewers MD, Knoell DL (2011) A comparison of zinc metabolism, inflammation, and disease severity in critically ill infected and noninfected adults early after intensive care unit admission. Am J Clin Nutr 93:1356–1364.

17. Lee YH, Bang ES, Lee JH, Lee JD, Kang DR, Hong J, Lee JM.. Serum concentrations of trace elements zinc, copper, selenium, and manganese in critically ill patients. Biological trace element research. 2019;188(2), 316–325.

18. Reeves PG, O’Dell BL. An experimental study of the effect of zinc on the activity of angiotensin converting enzyme in serum. Clin Che. 1985;31(4):581–4.

19. Li MY, Li L, Zhang Y, Wang XS. Expression of the SARS-CoV-2 cell receptor gene ACE2 in a wide variety of human tissues. Infect Dis Povert. 2020;9(1); 45

20. Devaux CA, Rolain JM, Raoult D. ACE2 receptor polymorphism: susceptibility to SARS-CoV-2, hypertension, multi-organ failure, and COVID-19 disease outcome. J Microbiol Immunol Infec. 2020;53(3):425–35,

21. Zoidis E, Seremelis I, Kontopoulos N, Danezis GP. Selenium-dependent antioxidant enzymes: Actions and properties of selenoproteins. Antioxidants, 2018; 7(5), 66.

22. Forceville X, Vitoux D, Gauzit R, Combes A, Lahilaire P, Chappuis P. Selenium, systemic immune response syndrome, sepsis, and outcome in critically ill patients. Crit. Care Med. 1998, 26, 1536–1544.

23. Braunstein M, Kusmenkov T, Zuck C,. Angstwurm M, Becker NP, Bocker W, Schomburg L,Bogner-Flatz V. Selenium and selenoprotein p deficiency correlates with complications and adverse outcome after major trauma. Shock 2020, 53, 63–70

24. Manzanares W, Langlois PL, Heyland DK. Pharmaconutrition with selenium in critically ill patients:What do we know? Nutr. Clin. Pract. 2015, 30, 34–43.

25. Maggini S, Pierre A, Calder PC. Immune Function and Micronutrient Requirements Change over the Life Course. Nutrients 2018, 10, 1531.

26. Saeed F, Nadeem M, Ahmed RS. Tahir Nadeem M, Arshad MS, Ullah A. Studying the impact of nutritional immunology underlying the modulation of immune responses by nutritional compounds—A review. Food Agric. Immunol. 2016, 27, 205–229.

27. Raha S, Mallick R, Basak S, Duttaroy AK. Is copper beneficial for COVID-19 patients?. Medical Hypotheses. 2020;109814.

